# Prevalence and correlates of anxiety and depression among female sex workers in Dire Dawa city Administration, Eastern Ethiopia

**DOI:** 10.1101/2023.02.03.23285415

**Authors:** Yassin Mohammed Yesuf, Dawit Mekonnen, Hareg Teklu, Sebsibew Atikaw Kebede

**Affiliations:** Department of Psychology, University of Gondar, Gondar, Ethiopia; School of Psychology, Addis Ababa University, Addis Ababa, Ethiopia; Department of Psychology, Dire Dawa University, Dire Dawa, Ethiopia; Department of Mathematics, University of Gondar, Gondar Ethiopia

**Keywords:** Anxiety, depression, sex work, female sex workers, Eastern Ethiopia, Dire Dawa city administration

## Abstract

**Background:** The mental health states of Female Sex Workers (FSWs) are least explored. The present study examined the levels of anxiety and depression as well as associated factors among FSWs in Dire Dawa city.

**Methods:** Cross-sectional explanatory sequential mixed-method research design was used. Snowball technique was used to recruit respondents. Beck Anxiety Inventory and Patient Health Questionnaire were used to measure anxiety and depression, respectively. Valid questionnaires were collected from 292 respondents (81% response rate) and interviews were made with six FSWs. Data collected through the questionnaire were analyzed using frequency, percentage, independent sample t-test, ANOVA, Pearson correlation and multiple linear regression while thematic analysis was used to analyze the qualitative data.

**Results:** The prevalence rates of anxiety and depression were 43.5% and 69.8%, respectively. Psychosocial and demographic variables were important correlates explaining 56% (F=43.324, p=<0.01, R2=.557) and 23% (F=10.319, p<0.01, R^2^=.228) of the variations in anxiety and depression, respectively. Specifically, *Khat* use (β= .371, t=8.744, p<0.01), violence (β= .272, t= 6.521, p<0.01), stigma (β= .258, t= 5.427, p<0.01), tobacco use (β=.200, t= 3.837, p<0.01) and alcohol use (β= -.136, t= -3.327, p<0.01) were significantly correlated with anxiety. Besides, violence (β= .298, t= 5.439, p<0.01), alcohol use (β= -.162, t= 3.011, p<0.01), stigma (β= .160, t= 2.549, p<0.05), and *Khat* use (β= .151, t= 2.915, p<0.01) were significantly associated with depression. The qualitative findings substantiated the survey results.

**Conclusion:** From the findings it is concluded that FSWs in Dire Dawa city were experiencing higher levels of anxiety and depression where psychosocial were important correlates of FSWs’ mental health. Therefore, organizations that are working with and/or concerned about FSWs need to provide evidence-based mental health and psychosocial interventions.

## Introduction

Mental disorders are among the leading causes of ill health and disability worldwide. In 2020 alone, there were 246 million people affected by depression and 341 million people were affected by anxiety disorders (1). In Ethiopia mental illnesses account for 11% of the total disease burden and the disability associated with mental illness is high (2). The total prevalence rate of common mental illnesses in Ethiopia is 21.8% (3).

Although no population group is immune from mental illnesses, some population groups are at higher risk of developing mental illnesses than others. Female Sex Workers (FSWs) are among the highly affected segments of the population in this regard. To put it in perspective, in a systematic review of studies on the mental health states of FSWs in low and middle income countries (LMICs), the pooled prevalence rate of depression was 41.8%, that of anxiety was 21.0%, PTSD was 19.7%, psychological distress was 40.8%, recent suicide ideation was 22.8% and recent suicide attempt was 6.3% (4).

The rates of mental illness prevalence found among FSWs were significantly higher than the general population. For example, in a study conducted in South Africa it was revealed that the level of anxiety among FSWs was 78% and that of depression was 80%. The study depicted that the rates among the FSWs were almost four times higher than the rates of anxiety and depression in the general population (5).

The high rates of mental illness among FSWs were attributed to a number of psychosocial variables. Different types of violence (6,5,7,4,8–11), stigma associated with sex work (5,12), substance use (4,6,13,8), STDs (sexually transmitted diseases) (6,4), inconsistent condom use (4,7), and poverty (6,8) were some of the factors reported to increase the risks of mental health problems among FSWs.

Although findings were inconsistent, a number of socio-demographic variables were also associated the high prevalence of mental health problems among FSWs. Age of the FSWs is a good example here. It was found to be associated with FSWs’ depression level in Uganda (10) and South Africa (14) while another study in South Africa found no association between age and anxiety and depression levels of the FSWs (5).

While these are the facts on the ground, studies over the mental health states of FSWs are limited (13) and are very limited in lower and middle income countries (8,10). And this is also the case in Ethiopia.

In Ethiopia there are studies that portrayed the prevalence of violence, stigma, substance use, unsafe sex, and sexual transmitted diseases among FSWs. Particularly studies among FSWs reported higher rates of work-related violence and alcohol abuse (Mooney et al., 2013), sexual violence (15), physical abuse, rape (16), stigma and discrimination (17,18), heavy episodic drinking (19) substance abuse and inconsistent use condoms (20), low rates of HIV preventive practices (21) and higher rates of STDs (22), including higher rates of HIV prevalence (23).

As described earlier, studies here and there depicted that the presences of these variables among FSWs were associated with elevated mental health problems. Thus, it can reasonably be argued that FSWs in Ethiopia are at increased risks of developing mental health problems. However, studies on the prevalence rates of mental health problems among FSWs in Ethiopia are scant while the available studies portrayed higher rates. A typical example here is a study that examined the depression level of rural-urban migrants in seven cities in Ethiopia. The study categorized the levels of depression based on respondents’ occupational status and found that the prevalence of depression was significantly higher among FSWs than the participants from other professions. Specifically, the study depicted that 26.9% of the FSWs experienced moderate depression while 9.8% of them experienced severe depression (24).

Likewise, there were two studies in Addis Ababa that examined the mental health states of street-based FSWs. The first study assessed the mental health of street-based FSWs using the mental health inventory (MHI-38) and found psychological distress among 35.4% of the sex workers (25). The second study examined PTSD, depression and substance abuse levels of street based FSWs. In the study it was reported that 31% of the FSWs experienced PTSD, around 60% of them experienced depression (mild, moderate and severe) and 61.5% of them were substance abusers (26).

These studies focused mainly on street-based FSWs, leaving aside FSWs in other work settings (home-based FSWs and those working in bars/hotels). However, the rates of mental health problems reported from the studies deserves due attention from researchers, practitioners and policy makers.

In Dire Dawa city administration, the present study area, there is a dearth of literature regarding the mental health states of FSWs even though there are studies which reported prevalence of higher rates of abuse/violence, substance abuse, and unsafe sex among the FSWs. For example, a study revealed that Hotel and Bar owners forced FSWs to drink alcohol and to have sex with customers for low prices, Bajaj drivers raped them while friends and intimate partner requested money to satisfy their addiction to *Khat* and alcohol (27). Another study reported inconsistent condom use, unsafe sex, fear of contracting HIV/STDs and perceptions of hazardous work environment by FSWs (28). Still another study depicted higher rates of life time substance use, current substance use, substance abuse and inconsistent condom use among FSWs (29).

These kinds of work related violence, inconsistent condom use, forced substance use and substance abuse would increase the vulnerabilities of FSWs to different mental health problems. Nevertheless, there is no available literature that indicated the prevalence of mental health problems among FSWs in the study area, at least to the knowledge of the current researchers. The present study, therefore, tried to fill the research gaps by assessing the prevalence and correlates of anxiety and depression among FSWs in Dire Dawa City Administration. Findings of the present study would be of great help for government and non-government organizations, practitioners, policy makers and civic societies who are working or at least concerned about FSWs. This is due to the fact that the results showed potential areas for interventions to improve the health of FSWs.

## Materials and Methods

### Research design

The study employed mixed-method research design. Of the available mixed-methods designs, explanatory sequential mixed methods design was employed where quantitative data was collected first and qualitative data was collected later to substantiate the findings from the quantitative data (30). In terms of the timing of data collection, the study employed cross sectional research design. Based on the analytic rigor, the study employed both descriptive and explanatory research designs. It is descriptive in that it summarized and described the anxiety and depression levels of the respondents as well as their socio-demographic characteristics. It is explanatory because it tests the association that exists between anxiety and depression and the predictor variables.

### Research setting

The study was conducted among FSWs in Dire Dawa City Administration which is found in Eastern part of Ethiopia 515 km away from the capital Addis Ababa. After Addis Ababa, Dire Dawa is Ethiopia’s second-largest urban area. According to 2007 census, the total population of Dire Dawa was 341,834, of whom 170,461 were women. Based on the census results 233,224 or 68.23% of the population were urban inhabitants which makes Dire Dawa the second most populous city in Ethiopia. The city has 9 urban and 38 rural Kebeles. In 2022 the total population of Dire Dawa city is projected to be 536,000, of whom 270, 000 will be women (31). Dire Dawa city is one of the major routes in the region for international migration. There are well-established ways that links Dire Dawa to Gulf States via Djibouti. Its strategic locations make Dire Dawa attractive for migration where most migrants work as sex workers and other labor activities in the courses of their migration.

### Population, Sample size and Sampling techniques

In this study, the target population was active FSWs who were living in Dire Dawa. The actual number of FSWs in Dire Dawa is unknown but years ago the number of sex workers who live in Dire Dawa city were estimated to be 2,254 (27). Hence, this number is used as the target population to decide on the sample size of the present study.

Since the target population is finite (known), we have used the Cochran (1977) formula to determine the sample size of this study. According to Cochran the ideal sample size for our population (2,254) is 384. The formula for definite population is n= n0/1+(n0-1)/N where n=sample size, n0=ideal sample size proposed and N=the population size. The calculation produced a sample size of 344. And we assumed a 5% non-response rate (17 in numbers) which makes the sample size of the present study to be 361.

When conducting research on and with FSWs, certain methodological issues arise that are related to the stigmatized nature of the work. As a result finding representative sample through randomization might not be practical, particularly when they are working in/from home settings. Therefore, during data collection we used snowball sampling techniques to recruit our sample.

In practice we started the data collection from FSWs that was purposefully selected by experts from Women, Children, Youth and Social Affairs Office (WCYSAO) from the different type of sex workers (based on how and where they meet their customers). After the data collectors meet with a FSW, the FSW was requested to facilitate contacts with other FSWs. During the recruitment of FSWs from bars/hotels the number of FSWs was first estimated first, then (a) all the FSWs were contacted when their number was estimated small or (b) 3-5 FSWs were contacted randomly if the number of the FSWs were estimated to be high. Take-all snowball technique is employed to select FSWs who work in the street, in their own home and in the clients’ house.

### Instruments

In the present study two instruments, i.e. questionnaire and interview were employed.

#### Questionnaire

The questionnaire is composed of four sections: socio-demographic measure, psychosocial measure, anxiety measure and depression measure.

##### Socio-demographic Measure

consists of items related to respondents’ characteristics (age, educational level, marital status, place of residence, having children, income level, types of sex work and years of service).

##### Psychosocial Measure

This measure includes tools to assess internalized stigma, violence and substance use among respondents. For the sake of examining violence among the FSWs, Violence Against Women instrument (VAWI) developed by WHO in 2005 was employed. VAWI is a 10 item tool used to examine exposure to sexual, physical and psychological violence. A guide on how to use was developed by WHO (32) and a modified version of the instrument was used here. The Alcohol, Smoking and Substance Involvement Screening Test (ASSIST V3.0) developed by WHO was used to identify substance use (WHO, 2002). Since ASSIST is a complex tool a modified version of it was used to gauge in to respondents’ use of tobacco, alcohol, *Khat* and marijuana use. There is no sex work-related stigma assessment method that measures all stigma characteristics and thus self-developed tool was used to measure stigma. The items were modified from The Internalized Stigma of Mental Illness Scale-9 (ISMI-9) developed by (33). Participants replied to five questions that addressed various forms of stigma experienced by sex workers (e.g. I am isolated from my friends because I am a sex worker). Likert rating scale was used that ranges from 0 (*Strongly disagree*) to 3 (*Strongly agree*). Total scores were computed where higher score indicates higher internalized stigma.

##### Anxiety Measure

The Beck Anxiety Inventory (BAI) was used to measure anxiety. BAI is a 21-item self-report instrument that assesses the severity of anxiety in adults and adolescents (Beck, Epstein, Brown, & Steer, 1988). Each item of the BAI describes a common symptom of anxiety. Respondents were instructed to report how bothersome each symptom has been over the past month using a 4 point Likert scale ranging from Not at all (0) to Mildly-but it don’t bother me much (1), Moderately-it was not pleasant at times (2) and Severely-it bothered me a lot (3). All of the items were summed to provide a total score. Scores can range from 0 to 63 where scores from 0 to 21 indicate mild anxiety, scores from 22 to 35 indicate moderate anxiety and scores above 35 indicate severe anxiety. The BAI has demonstrated good psychometric properties. In the initial psychometric study, test retest reliability was good (r= .75), and internal consistency was high (α=.92) (Beck, Epstein, Brown, & Steer, 1988).

##### Depression Measure

To measuring the severity of depression over the past two week Patient Health Questionnaire (PHQ-9) was used. It assessed how often have the respondents been bothered by any of the listed 9 problems using a 4 point Likert scale (Not at all (0), several days (1), More than half the days (2), and Nearly every day (3). All of the items were added together to get a total score that ranges between 0 and 27. The composite scores were then categorized in to levels of depression as: no depression (0–4), mild depression (5–9), moderate depression (10–14), moderately severe depression (15–19) and severe depression (20–27)

#### Interview guide

In this study, researcher-developed semi-structured in-depth interview guide was employed to gather data from FSWs selected using snowball technique. These in-depth interviews focused on illegal drugs, experiences of violence, stigma and other work related experience of FSWs that are related to mental health problems.

### Data collection Procedure and Ethical considerations

The study was approved by Gondar University, department of psychology review board. Due to the stigmatized, marginalized and often criminalized nature of the sex industry, it was important that sex workers were assured of confidentiality and privacy as this is the corner stone of a trusting relationship. To maintain confidentiality of the information provided by the respondents, the respondents were instructed not to write their name on the questionnaire, and assured that responses will be used solely for academic purpose and aggregate results will be reported.

The tools (the questionnaire and the interview guide) were first prepared in English. The research team together with a language expert translated the tools to Amharic. The tools were then back translated by psychologists and language experts who were not familiar with the purpose of the study. And minor differences in translations were resolved through a focus-group discussion.

Prior to the main data collection through the questionnaire, pilot test was conducted among 36 FSWs in Harar town (10% of the sample). This was done to check the reliability and applicability of the questionnaire developed. In terms of reliability the Cronbach alpha of the instruments range between .796 and .835 indicating acceptable to good reliability. With regard to applicability typing and grammatical errors were corrected; and some items (e.g. items related to cocaine use) were deleted based on the feedbacks from the respondents.

Data collectors were trained on data collection through questionnaire. The main data was collected with the support from the City Administration WCYSAO guided by the research team. The qualitative data was exclusively collected by the third researcher (HT).

### Data Analysis

Data collected through the questionnaire were analyzed using descriptive and inferential statistics. Frequencies and percentages were used to describe the profile of respondents and to describe respondents’ anxiety and depression level. Series of independent sample t-tests and ANOVA testes were used to look in to mean differences in respondents’ anxiety and depression level based on background variables. Pearson correlations were computed to check into the relationships between the criterion and the predictor variables. Multiple linear regression analyses were executed to determine the effects of predictor variables over respondents’ anxiety and depression. Thematic analysis was used to analyze the qualitative data.

## Results

Of the total 361 questionnaires distributed, 3 respondents failed to return the filled questionnaires. Out of 358 questionnaires collected from the respondents, 292 were complete and 66 were discarded due to high volume of missing values. This made the overall response rate to be 81%.

### Socio-demographic Characteristics of the Respondents

Respondents’ age, educational level, marital status, residence, income level, children, type of sex work and years of service were treated as the socio-demographic characteristics. Thus, the following table presents each characteristic described using frequency and percentage.

Table 1 showed that 52.1% of the FSWs don’t have children, 51% of them aged between 26 and 32 years, 51.4% of them attended elementary education and 51.7% of them earn more than 7000 birr per month.The Table also informs us that 45.5% of the respondents work in hotels and bars, 28.8% of them were divorced, 65.4% of them lived in rural areas and 67.8% of them stayed between 1 and 4 years as sex workers.

**Table 1.**
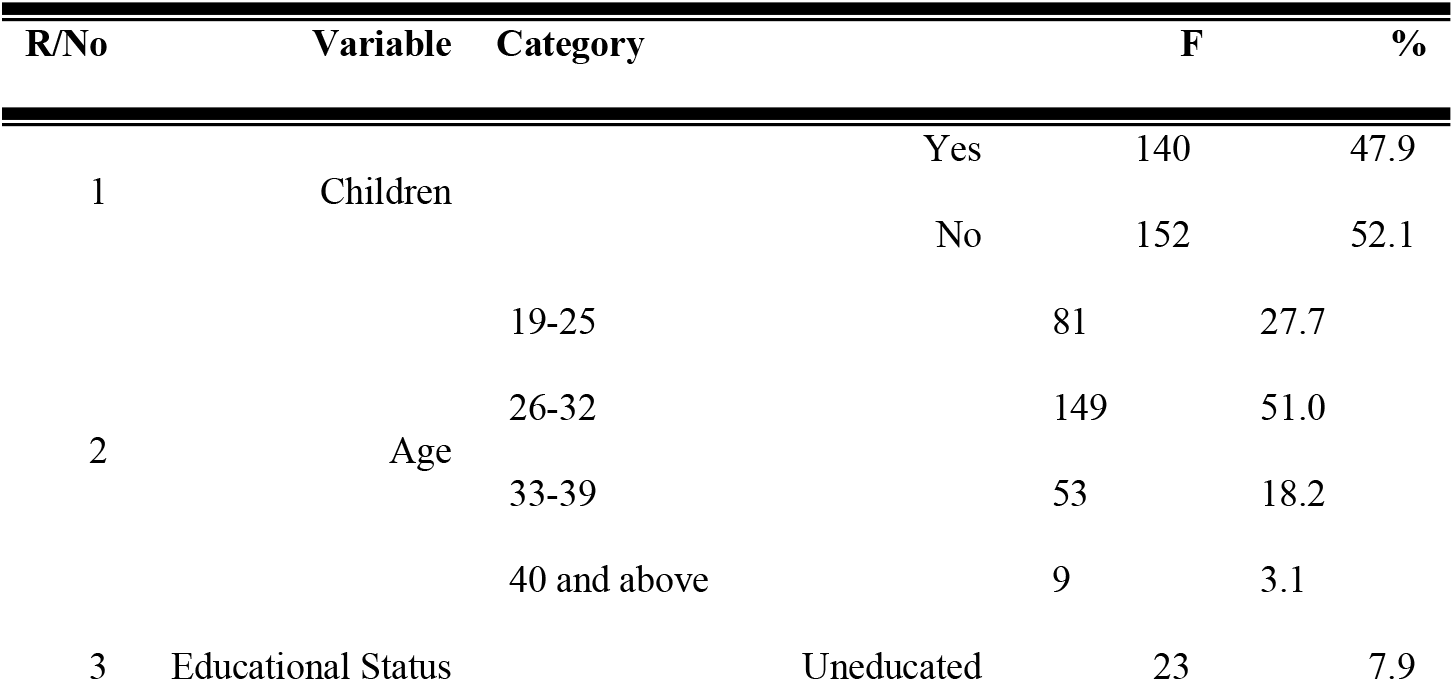
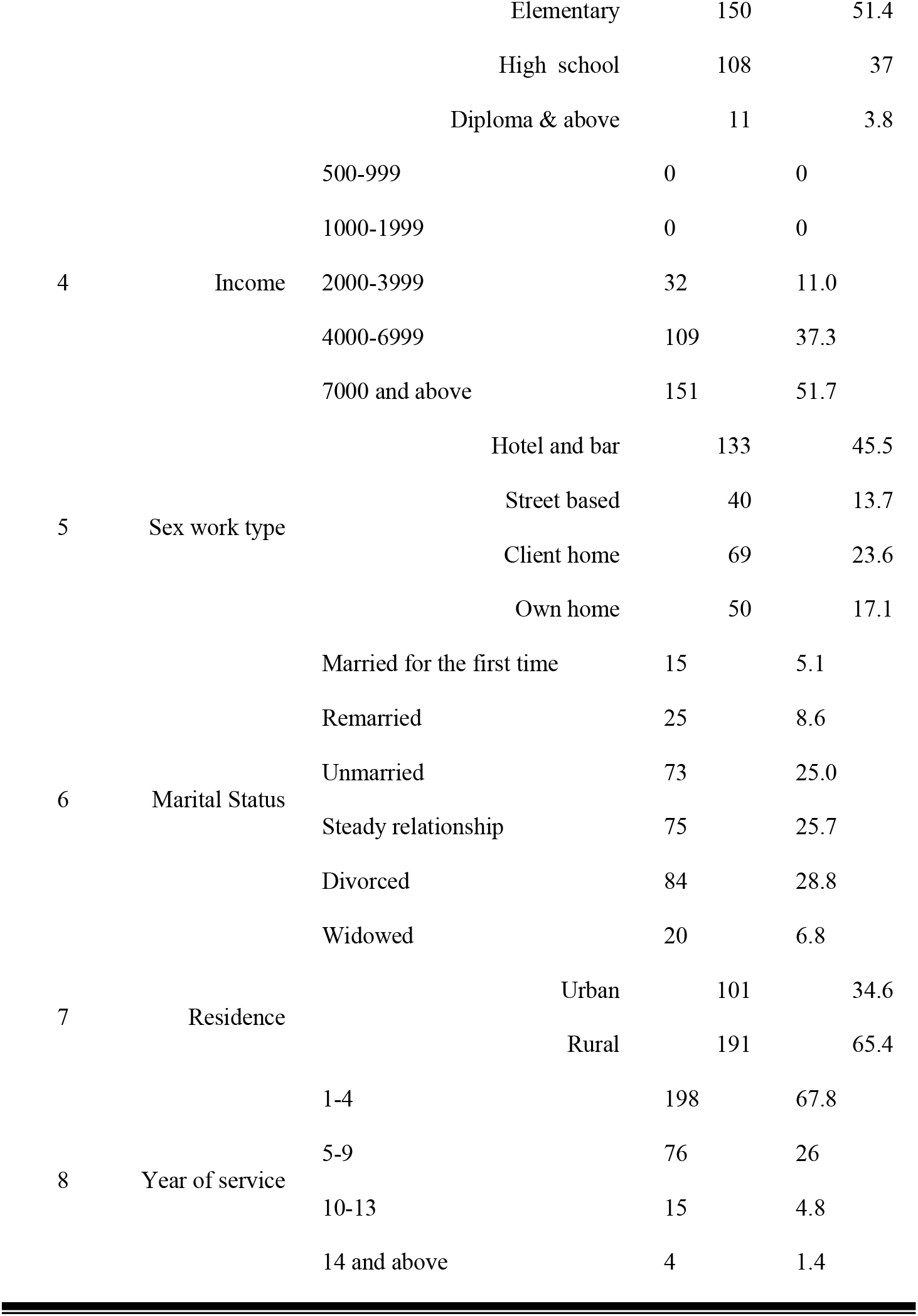
*Socio-demographic characteristics of the respondents* (N=292)

### The Prevalence of anxiety and depression among FSWs

As its first objective, this study examined the prevalence of anxiety and depression among FSWs. Frequency and percentage analysis were undertaken to achieve this objective. The following table depicted the results of the computations.

Table 2 presents the prevalence of depression and anxiety among FSWs who participated in this study. According to the table, 107 (36.6%) of the respondents have severe depression, 74 (25.3%) of them have moderate depression, 54 (18.5%) of them have mild depression, and 23 (7.9 %) of the respondents have moderately severe level of depression. The remaining 34 respondents (11.6%) have no depression. Hence, the majority of the respondents of the study had severe depression.

**Table 2.** *The Prevalence of depression and anxiety among FSWs* (N=292)

| **Variable** | **Category** | **N** | **%** |
| --- | --- | --- | --- |
| Depression | No depression | 34 | 11.6 |
|  | Mild depression | 54 | 18.5 |
|  | Moderate depression | 74 | 25.3 |
|  | Moderately severe depression | 23 | 7.9 |
|  | Severe depression | 107 | 36.6 |
| Anxiety | Mild Anxiety | 165 | 56.5 |
|  | Moderate Anxiety | 66 | 22.6 |
|  | Severe Anxiety | 61 | 20.9 |

Table 2 also demonstrated the prevalence of anxiety among FSWs. As can be inferred from the table, those participants who reported to have mild anxiety account for 56.5% of the respondents. Sixty six participants (22.6%) have moderate level of anxiety and 61 participants (20.9%) have severe anxiety level. Thus, the majority of participant in the study area were found to have mild level of anxiety.

### Mean differences in anxiety and depression

The other focus of the present study was examining significant mean difference in the level of depression and anxiety among respondents across socio-demographic variables. The following table presented results of the analysis of ANOVAs and independent sample t-tests that were used to examine mean differences in anxiety.

Table 3 depicted that there were no statistically significant differences in anxiety based on respondents’ socio-demographic characteristics. The only exception here is sex work type (F= 3.947, p<0.001). Post hoc comparisons using Tukey HSD test indicated that street sex workers (M= 27.50, SD= 9.821) have higher anxiety level than respondents who work in their home (M= 21.74, SD= 7.972).

**Table 3.**
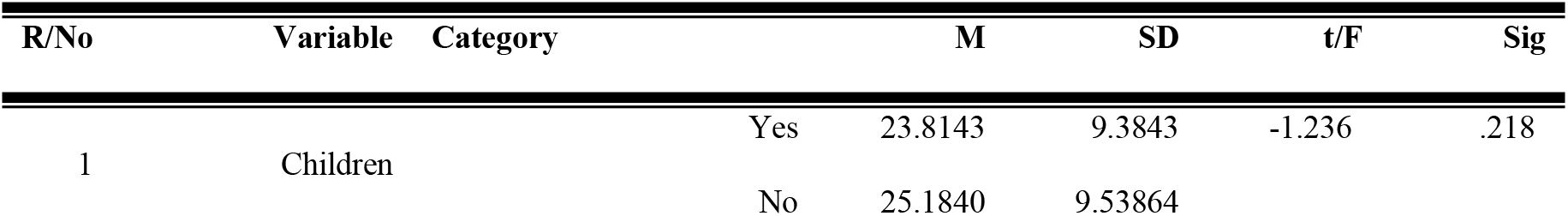
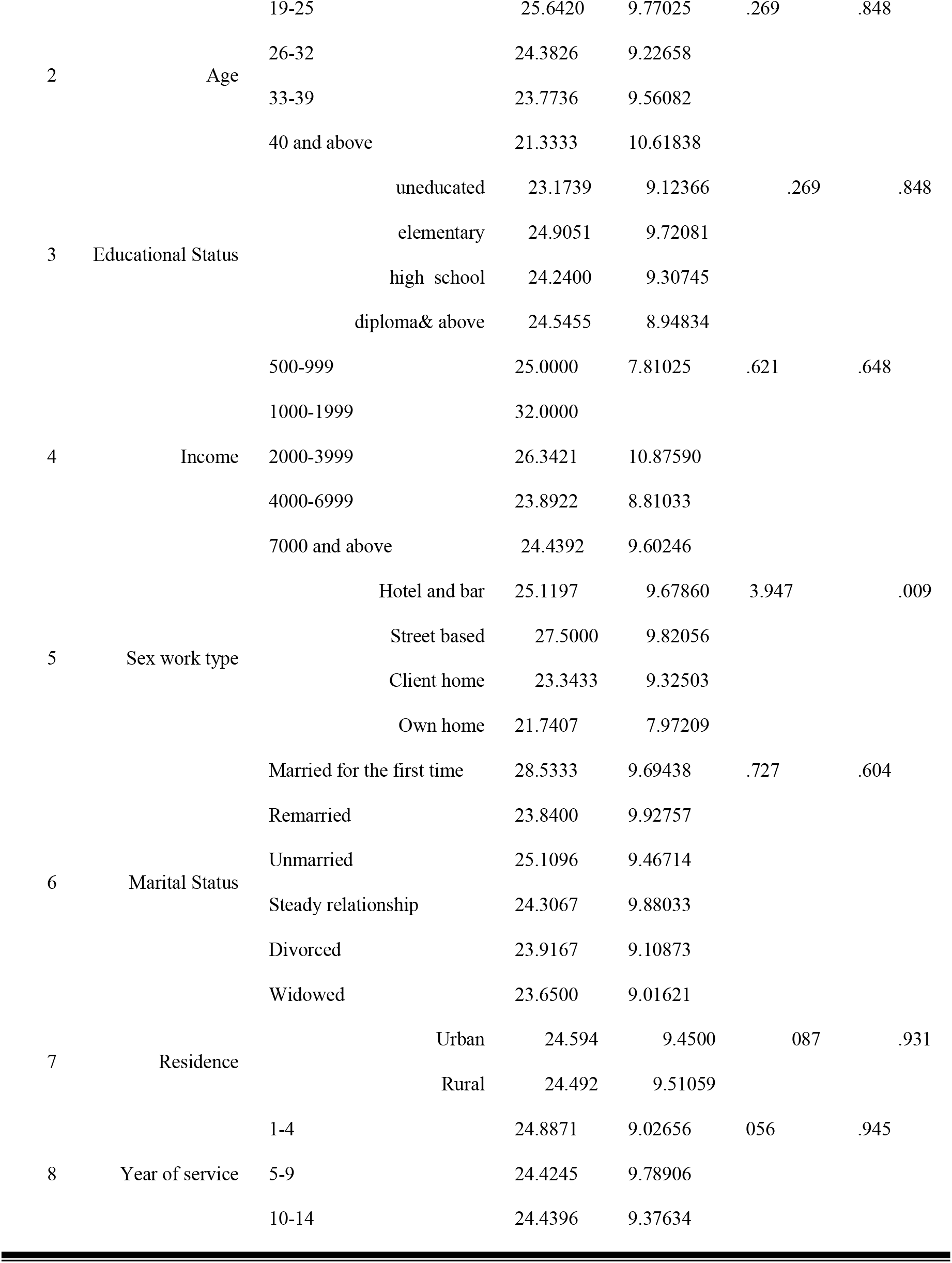
*Mean differences in anxiety across socio-demographic variables* (N=292)

The following table presents results of the analysis of ANOVA and independent sample t-test results conducted to look into mean differences in depression based on socio-demographic variables.

As can be seen from Table 4 there were no statistically significant differences in depression based on respondents’ socio-demographic characteristics.

**Table 4.**
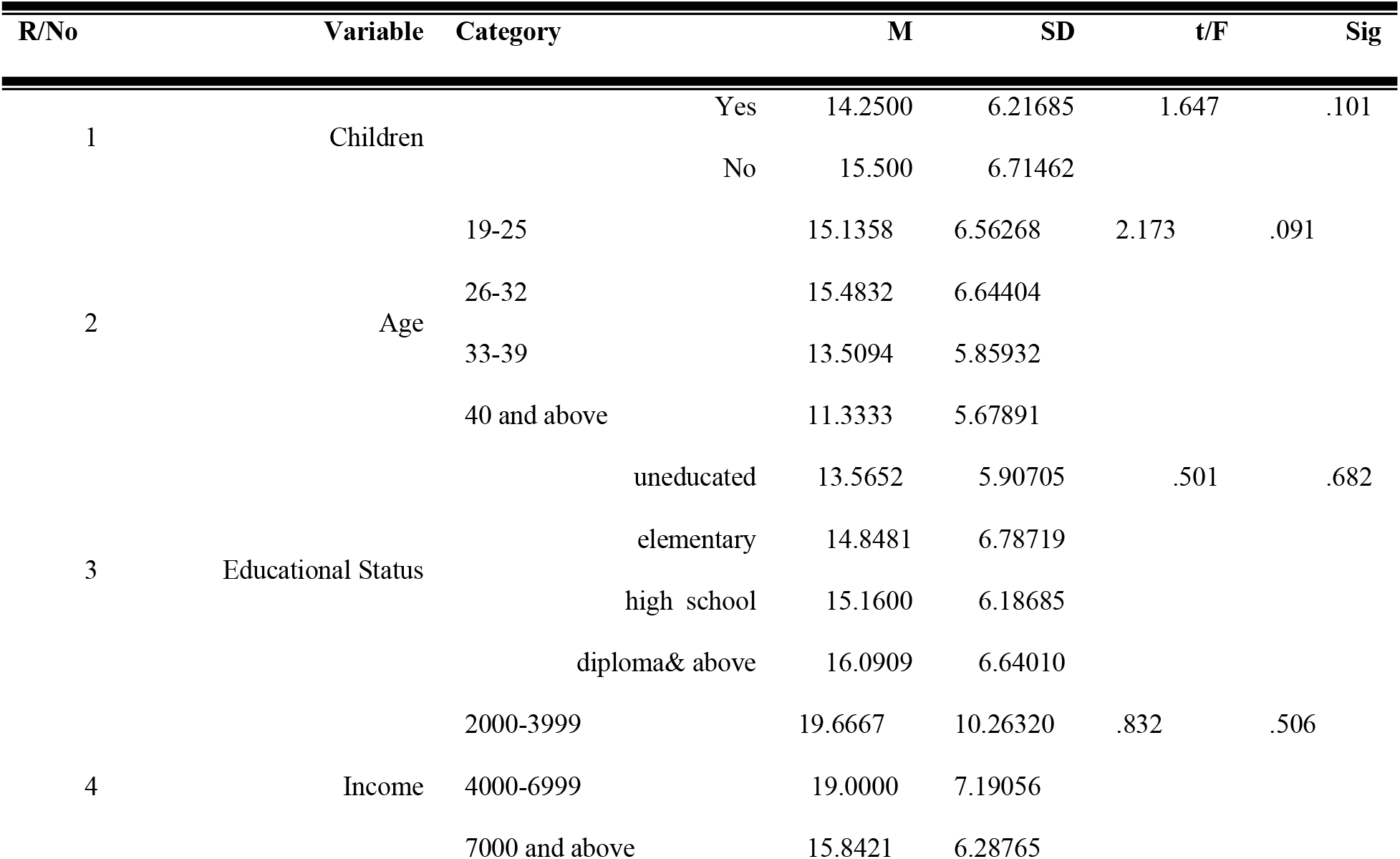
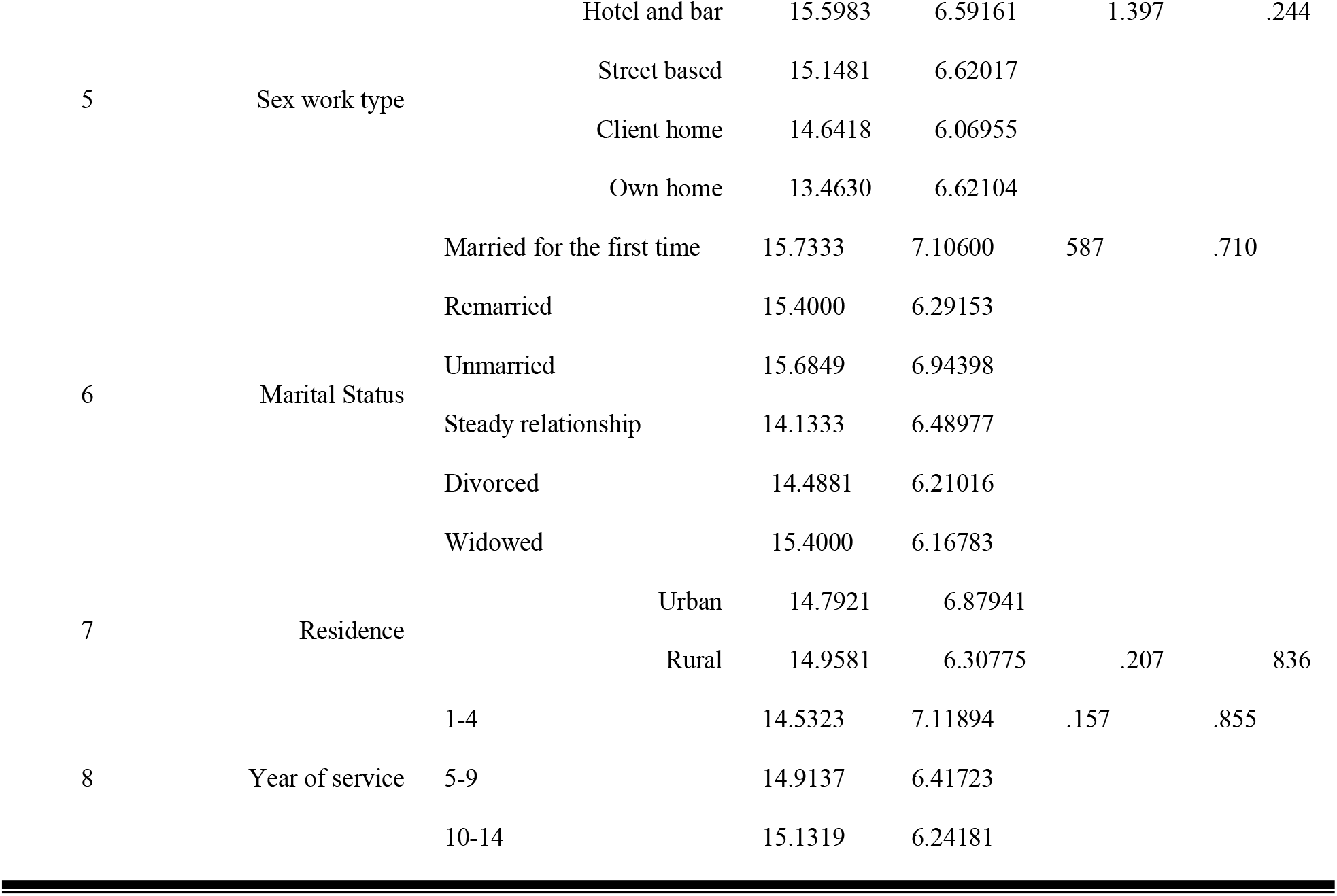
*Mean differences in depression across socio-demographic variables* (N=292)

### Correlates of anxiety and depression

In order to identify the predictor variables that influence respondents’ anxiety and depression, multiple linear regressions were computed. Preliminary assumption tests were conducted to check for normality, linearity, multicollinearity, outliers, homoscedasticity and independence of residuals. Results showed that there were no major violations of these assumptions.

Prior to the regression analysis, association among predictor variables (socio-demographic and psychosocial variables) and the criterion variables (anxiety and depression) were checked using Pearson’s correlation coefficient. Based on the results of the computations two demographic variables (educational status and service year) and the six psychosocial variables were found to have statistically significant relationship with both anxiety and depression. Thus, two regression models were computed taking those related variables as predictor variables and the results of the computations are presented in Table 5 below:

**Table 5.** *Linear regression analysis of independent variables to the dependent variables*

|  | **b** | **SEb** | **$\beta$** | **t** | **p** | **R<sup>2</sup>** | **F value** |
| --- | --- | --- | --- | --- | --- | --- | --- |
| **Model 1 Anxiety** |  |  |  |  |  | .557 | 43.324 ** |
| Violence | .604 | .093 | .272 | 6.521 | .000 |  |  |
| Tobacco | .266 | .069 | .200 | 3.837 | .000 |  |  |
| Alcohol | -.304 | .091 | -.136 | -3.327 | .000 |  |  |
| *Khat* | .522 | .060 | .371 | 8.744 | .001 |  |  |
| Marijuana | .036 | .049 | .041 | .748 | .455 |  |  |
| Stigma | .739 | .136 | .258 | .427 | .000 |  |  |
| Service year | .412 | .538 | .031 | .766 | .445 |  |  |
| Education | .490 | .565 | .035 | .866 | .387 |  |  |
| **Model 2 Depression** |  |  |  |  |  | .228 | 10.319** |
| Violence | .450 | .083 | .298 | 5.439 | .000 |  |  |
| Tobacco | -.003 | .062 | -.003 | -.048 | .962 |  |  |
| Alcohol | .246 | .082 | -.162 | 3.011 | .003 |  |  |
| *Khat* | .145 | .053 | .151 | 8.744 | .007 |  |  |
| Marijuana | .053 | .043 | .087 | 1.212 | .226 |  |  |
| Stigma | .310 | .122 | .160 | 2.549 | .011 |  |  |
| Service year | .461 | .481 | .052 | .958 | .339 |  |  |
| Education | .654 | .506 | .069 | 1.294 | .197 |  |  |

As can be seen from Table 5 the overall model fit in Model one was significant (F=43.324, p=<0.01, R^2^=.557) implying that 56% of the variations in anxiety were explained by the 8 predictor variables included in the model. Based on the standardized beta coefficient values the highest contribution in explaining anxiety came from *Khat* use (β= .371, t=8.744, p<0.01). That is followed by violence (β= .272, t= 6.521, p<0.01), stigma (β= .258, t= 5.427, p<0.01), tobacco use (β=.200, t= 3.837, p<0.01) and alcohol use (β= -.136, t= -3.327, p<0.01). From the t-test results of the beta coefficient it can be seen that service year, education and marijuana use were insignificant predictors of anxiety.

Model 2 in Table 5 depicted that when all predictor variables were considered together, they significantly predicted 23% of the variations in depression (F=10.319, p<0.01, R^2^=.228). The standardized beta coefficient values indicated that the highest contribution in explaining depression came from violence (β= .298, t= 5.439, p<0.01), followed by alcohol use (β= -.162, t= 3.011, p<0.01), stigma (β= .160, t= 2.549, p<0.05), and *Khat* use (β= .151, t= 2.915, p<0.01).From the t-test results of the beta coefficient values it can be seen that tobacco use, marijuana use, service year and educational status were insignificant predictors of FSWs’ depression.

### Qualitative Data

Thematic analysis was used to scrutinize the data collected through the in-depth interviews. The themes that emerged at first glance were the push and the retaining forces in the sex work. All the respondents mentioned that financial problems were the reasons behind entering in to the sex work business. Freedom of the work and relatively higher earnings were stated as retaining factors in the business. An extract from a respondent will show how financial strain and gain were the pushing and retaining factor in the sex work business. She utters:

> *“Currently, I am living with my two children. Before I started this business I was in a deprived situation and I didn’t have money to fulfill my children’s basic needs. Also when I compared this work with other [work type]in terms of its income, this work is better because I could get better money in short period time with low educational status and no vocational training”* (Interviewee 2)

Moreover, majority of the participants had restricted social life, which were only limited to other sex workers, owners’ of brothel (‘*Baluka’* as they call it) or a man that serve as a body guard. Their social life is crimpled because of the work related stigma they have experienced. One of the study participants shared her experience as follows:

> *“My social interaction is not good with other people because I heard some gossip about my work. I tried to hide my work but through time it becomes noticeable. When people know about my work they change their greetings and their interaction with me. If I have some contact and let’s say take their children, they don’t want such kinds of interaction you know why they are Yebet lij [descent girl who never leaves her house] and I am a sex worker”* (Interviewee 4).

When the interviewees were asked about their experience of consuming substances, the majority of the participants replied that they chew *khat* daily, drink alcohol and take *Shisha*. But they don’t actually want to talk about illicit drugs. Thirty years old FSW described her substance use and its purpose as follow:

> *“I take different drugs such as: Khat, alcohol, cigarette and Gaya [a term they use interchangeably with Shisha]. To be honest I used Eka [a term for marijuana]. Sometimes I chew Khat and take Shisha for I can’t do anything if I don’t get them. Unless I get Khat and Gaya at my usually time, I show aggressive behavior and I will have Dukak [low mood]. I spent all of my time in my bed”* (Interviewee 6).

Regarding experience of violence, majority of the interviewed participants replied that they experienced different forms of violence where sexual violence was typical one. A 29 years old sex worker expressed her experience as follow:

> *“I face different violence from customers because of sexual intercourse type [sex position]. Some want to have sex by watching pornography and experiment on me as a toy, sometimes they come with other sex workers after they take different drugs that will help them to prolong their duration of sexual intercourse, it is difficult to handle this. Moreover, I don’t know how much it is common but some costumers asked me totally unusual things that they may want beating and assailing during sexual intercourse”* (Interviewee 1).

When the respondents were asked about their thoughts towards their work, most of the respondents explained that they feel awful. Some pointed out the psychological distress they undergone through. For example one of the study participants said:

> *“Totally I am not comfortable selling my bodies for money…*.*I assume that there is no dignity, self-respect… sometimes I get nervous. Always I feel inferior when I compared my current situation with my situation at my early age and my class mates”* (Interviewee 3).

Generally speaking the interview participants indicated the presence of stigma, sexual violence, drug use and psychological distress in their work environment.

## Discussion

To the knowledge of the present researchers this study is the first of its kind in Ethiopia in that it portrayed the prevalence and correlates of anxiety and depression among FSWs using mixed methods approach. Our work contributes to the little body of literature highlighting the vulnerability of FSWs to mental health issues.

Our first objective of the present study was assessing the prevalence of anxiety and depression among FSWs in Dire Dawa city. In the present study the overall prevalence of depression (moderate, moderately severe and severe) was 69.8% while the overall prevalence of anxiety (moderate and severe) was 43.5%.

In the global literature different anxiety and depression prevalence rates were reported among FSWs where some were higher, others comparable and some others were lower than the rate found in our study. For instance, the rates of depression levels range from 9% among FSWs in China (Tam et al., 2022) to 80.9% among FSWs in South Africa (5).

The overall rate of depression found in our study among FSWs in Dire Dawa city is significantly higher than the rate reported in the general population (11%) in Ethiopia (34). However, the rate in this study was lower than what was reported in South Africa and the Dominican Republic. In the study in South Africa (5) depression was reported among 80.9% of the female sex workers included while in the study in the Dominican Republic depression was prevalent among 70.2% of the respondents (12). Methodological differences were the possible explanation for the differences in the findings. For example, the South African study used PHQ-9 with cut off score of 4 while we categorized a respondent as depressed when she scored at least 10 (the cut off score to be categorized a respondent in moderate depression).

There were also studies reported comparable depression prevalence rates to ours. The study in Soweto, South Africa, revealed a prevalence of 68.7% depression among FSWs (35), and the study among FSWs working on the streets of Addis Ababa found a rate of 59% (26).

On the other hand, lower rates of depression prevalence than what was found in our study were reported from studies in other countries. In the study in Kenya (9) 56.8% of the respondents were found to have depression. In the study in South Africa a 52.7% prevalence rate of depression was reported (14). Similarly, a study in China showed that 52.4% of the FSWs have the potential to develop depression (36), the study in Northern Uganda reported a prevalence rate of 50.4% depressive symptoms among FSWs (10) and in a study in Cameron a depression prevalence rate of 49.8% was reported. Mythological differences and characteristics of the participants included could explain the differences in findings. For example, the study in China used GHQ-12 (36) to examine depression while our study used PHQ-9. Likewise, the study in Uganda was conducted in post-conflict area (10) while our study was conducted in a relatively peaceful area.

Significantly lower rates of depression in depression were reported from a study in China (37) and in Malawi (38). The study in Malawi reported a 9% prevalence rate of probable depression while the study in China reported depression among 8% of the FSWs. The tools employed to measure depression could possibly explain the differences in findings. The study in China employed the Chinese Version of Center for Epidemiologic Studies Depression Scale Revised (37). On the other hand, the Malawi study employed PHQ-9 to measure depression that is similar to our study. However, the authors acknowledged that PHQ-9 was not validated in Malawi (38). With regard to the prevalence of anxiety the present study found a prevalence rate of 43.5%. A comparable anxiety prevalence rate was reported from the study in Kenya (9) that reported anxiety among 39.1% of the FSWs. Contrary to these findings, another study in Kenya (8) reported an anxiety prevalence among 11% of the FSWs. Coupled with this the prevalence rate of anxiety found among FSWs in our study is significantly higher than the 21% prevalence rate reported from a systematic review conducted in LMIC (4). The rate found in our study was also higher than the 19.9% life time anxiety prevalence rate found among sex workers in Canada (13).

In terms of psychosocial variables associated with anxiety and depression, the present study depicted that violence, stigma and substance use were important correlates of the anxiety and depression levels of FSWs. Findings of the qualitative data have also substantiated findings of the quantitative data by highlighted the presence of substance use, stigma, violence and psychological distress.

In line with our findings, studies conducted here and there consistently showed that violence (4–11) and stigma (5,12) were significantly associated with FSWs’ anxiety and depression levels.

The literature over the association between substance use and FSWs’ anxiety and depression consistently depicted significant associations (4,6,8,13). Likewise, findings of the present study confirmed these associations. Of the substance use variables considered in the present study, alcohol use and *Khat* use were associated with both anxiety and depression. Tobacco use was associated with anxiety but not depression while Marijuana use was not associated with both anxiety and depression. These findings implied that substance use is an important correlate with FSWs’ anxiety and depression level although the correlations were dependent on the type of substance consumed.

With regard to socio-demographic variables, the present study found insignificant association between socio-demographic variables and the anxiety and depression levels of FSWs. Similar findings were reported from other studies. For instance, the study in South Africa (5) and the study in India (39) found no association between socio-demographic characteristics and the prevalence of depression and anxiety. On the other hand, others studies found association between socio-demographic variables and the prevalence of depression and anxiety. For example older age was associated with anxiety and depression in Nairobi Kenya (8) and with depression in South Africa (14). Similarly, age at first sex work was associated with depression in the study in Sewoto South Africa (35) while age at first sex work and education were associated with depression in Malawi (38). The differences in findings were attributed to methodological differences and characteristics of the FSWs included in the studies. These findings call up on further studies that examine the association between the mental health and demographic characteristics of FSWs.

The study is not without limitations. Its cross-sectional nature, the sampling techniques employed and the small number of interview participants were acknowledged as the major limitations of the study.

## Conclusions

Based on the results of the study it can be concluded that FSWs in Dire Dawa city experienced higher levels of anxiety and depression. At the same time it can be concluded that psychosocial variables were important correlates of the mental health problems of FSWs in the city. Therefore, it is recommended that government organizations (including the ministry of women and social affairs and the ministry of health), NGO, civil societies and others working with and/or concerned about FSWs need to provide mental health and psychosocial interventions, including counseling services, alongside existing interventions. Needless to say, the study is delimited to FSWs in Dire Dawa city. Thus, it is imperative to conduct larger study, possibly an implementation study, among nationally representative FSWs to deliver evidence-based interventions.

Comparing the findings of the study with other studies was challenging because of the variations in the tools used to measure anxiety and depression. Adapting or developing universal tools that measure anxiety and depression among FSWs will help to see the true difference between studies. Adapting/developing tools is, therefore, one of the future research areas. Besides, findings over the effects of demographic variables on FSWs mental health problems are still inconclusive thereby require larger global studies.

## Data Availability

The data underlying the results presented in the study are available from the corresponding author (YM) upon reasonable request.

## Acknowledgements

We acknowledge participants of the study and all the employees of the Dire Dawa city Administration Women, Children, Youth and Social Affairs Office for their contribution towards the smooth completion of the study.

